# Performance of Repeat BinaxNOW SARS-CoV-2 Antigen Testing in a Community Setting, Wisconsin, November-December 2020

**DOI:** 10.1101/2021.04.05.21254834

**Authors:** Melisa M. Shah, Phillip P. Salvatore, Laura Ford, Emiko Kamitani, Melissa J. Whaley, Kaitlin Mitchell, Dustin W. Currie, Clint N. Morgan, Hannah E. Segaloff, Shirley Lecher, Tarah Somers, Miriam E. Van Dyke, John Paul Bigouette, Augustina Delaney, Juliana DaSilva, Michelle O’Hegarty, Lauren Boyle-Estheimer, Fatima Abdirizak, Sandor E. Karpathy, Jennifer Meece, Lynn Ivanic, Kimberly Goffard, Doug Gieryn, Alana Sterkel, Allen Bateman, Juliana Kahrs, Kimberly Langolf, Tara Zochert, Nancy W. Knight, Christopher H. Hsu, Hannah L. Kirking, Jacqueline E. Tate

**Affiliations:** Coronavirus Disease 2019 (COVID-19) Response Team, Centers for Disease Control and Prevention, Atlanta, Georgia, USA; Epidemic Intelligence Service, Centers for Disease Control and Prevention, Atlanta, Georgia, USA; Laboratory Leadership Service, Centers for Disease Control and Prevention, Atlanta, Georgia, USA; Wisconsin Department of Health Services, Madison, Wisconsin, USA; Marshfield Clinic Research Institute, Marshfield, Wisconsin, USA; Winnebago County Health Department, Oshkosh, Wisconsin, USA; Wisconsin State Laboratory of Hygiene, Madison, Wisconsin, USA; University of Wisconsin-Oshkosh, Oshkosh, Wisconsin, USA

## Abstract

Repeating the BinaxNOW antigen test for SARS-CoV-2 by two groups of readers within 30 minutes resulted in high concordance (98.9%) in 2,110 encounters. BinaxNOW test sensitivity was 77.2% (258/334) compared to real-time reverse transcription-polymerase chain reaction. Repeating antigen testing on the same day did not significantly improve test sensitivity while specificity remained high.

## Manuscript

Strategies to curb the current coronavirus disease 19 (COVID-19) pandemic are increasingly reliant on antigen-based diagnostics because of low cost, availability, and rapid turn-around. Abbott’s BinaxNOW COVID-19 Ag Card (here on referred to as BinaxNOW) is a lateral flow antigen test resulting in 15 minutes and was initially approved for use in symptomatic individuals [1]. Recent reports suggest lower sensitivity compared to real-time reverse transcription-polymerase chain reaction (RT-PCR) among asymptomatic individuals [2-5]. The ideal frequency of serial rapid tests and the role of confirmatory rapid testing at the same encounter is not well-described. To evaluate repeat antigen testing and test performance, this investigation embedded repeat BinaxNOW testing and simultaneous RT-PCR confirmation at a testing site where antigen tests were freely available to the public regardless of symptoms or exposures [6]. Concordance of repeat BinaxNOW testing as well as test performance in a community setting with high prevalence of SARS-CoV-2, the virus that causes COVID-19, is described.

## Methods

Participants were recruited from registrants at a community SARS-CoV-2 testing site in Oshkosh, Wisconsin, during an ongoing mass surge testing campaign. The surge testing campaign was sponsored by the Wisconsin Department of Health Services, University of Wisconsin, and the U.S. Department of Health and Human Services [6]. For this investigation, individuals were recruited to undergo two BinaxNOW tests and an RT-PCR test regardless of their symptom status or result of the initial BinaxNOW test. The first BinaxNOW test was done by trained staff from the routine testing site while the second BinaxNOW test was done by a separate group of trained CDC staff, both according to manufacturer’s instructions. All swabs were supervised, self-collected, and from the anterior nares. Participants completed a questionnaire on demographics, exposures, and symptoms. Approximately 30 minutes after the initial swab was taken, each participant provided documentation of their initial BinaxNOW test result and two additional self-collected swabs were taken by CDC staff. Participants were instructed to simultaneously insert one swab into the left nostril and one swab into the right nostril, rotate five times, swap nostrils, and rotate five times again. One swab was used to perform the second BinaxNOW test immediately, and the other swab was placed in viral transport medium and transported to the Marshfield Clinical Research Institute laboratory for RT-PCR testing within two to five days. BinaxNOW results were considered invalid/indeterminate if no lines were seen in the results window or if only the sample line was seen. A three viral target RT-PCR assay (S gene, N gene, Orf1Ab) for SARS-CoV-2 was conducted. Positive specimens were defined as having at least two targets with a threshold cycle (Ct) value _≤_37 per manufacturer’s instructions [7]. MagMAX Viral/Pathogen Nucleic Acid Isolation Kit (REF A48310) was used for RNA extraction. Specimens with inconclusive results (defined as one of three positive targets) were re-tested. Symptomatic participants were defined as reporting one or more of 15 symptom criteria at enrollment [8]. RT-PCR was the gold standard for defining antigen test performance. Ninety-five percent confidence intervals (CI) for sensitivity, specificity, positive predictive value (PPV), and negative predictive value (NPV) were calculated using the Clopper-Pearson method. The Mann-Whitney-U test was used to test rank differences for Ct values; chi-square tests were used to test for differences in categorical variables; and t-tests were used to test for differences in ages. Statistical significance was defined as p<0.05. Statistical analysis was performed using R 1.3.1056.

## Results

Between November 16, 2020, and December 15, 2020, data on 2,127 participant encounters were collected, capturing 22% of routine tests performed at the community testing site. Six inconclusive RT-PCR results, seven missing RT-PCR results, and four indeterminate BinaxNOW test results from separate encounters were excluded for a total of 2,110 participant encounters with all tests (two BinaxNOW and one RT-PCR) among 2,024 unique individuals. Children under the age of 18 years provided 10.7% (225/2,110) of specimens (Supplementary Table 1).

**Table 1:** Concordance of repeated Abbott’s BinaxNOW antigen testing (N=2,110) for SARS-CoV-2 and performance characteristics compared with real-time reverse transcription-polymerase chain reaction (RT-PCR)^a^ results stratified by symptom status at a community testing center – Oshkosh, Wisconsin, November 16 - December 15, 2020

|  | Category | Total N=2,110 | RT-PCR Positive | N Gene Threshold Cycle<sup>b</sup> |
| --- | --- | --- | --- | --- |
| **BinaxNOW Tests Concordant** | All Concordant Tests | 2,086 (98.9%) | 315/2,086 (15.1%) | 22.3 |
|  | Both Positive | 255 (12.1%) | 253/255 (99.2%) | 20.2 |
|  | Both Negative | 1,831 (86.8%) | 62/1,831 (3.4%) | 30.7 |
| **BinaxNOW Tests Discordant<sup>c</sup>** | All Discordant Tests | 24 (1.1%) | 19/24 (79.2%) | 26.7 |
|  | Positive → Negative | 10 (0.5%) | 5/10 (50.0%) | 28.7 |
|  | Negative → Positive | 14 (0.7%) | 14/14 (100.0%) | 26.0 |
|  | Sensitivity %<br>(n, 95%CI)<sup>d</sup> | Specificity %<br>(n, 95%CI) | PPV %<br>(n, 95%CI) | NPV %<br>(n, 95%CI) |
| All Participants, Initial BinaxNOW test, (n=2,110) | **77.2**<br>(258/334, 72.4–81.6) | **99.6**<br>(1,769/1,776, 99.2–99.8) | **97.4**<br>(258/265, 94.6–98.9) | **95.8**<br>(1,769/1,845, 94.9–96.7) |
| All Participants, Repeated<sup>e</sup> BinaxNOW test, (n=2,110) | **81.4**<br>(272/334, 76.8–85.5) | **99.6**<br>(1,769/1,776, 99.2–99.8) | **97.5**<br>(272/279, 94.9–99.0) | **96.6**<br>(1,769/1,831, 95.7–97.4) |
| Symptomatic Participants<sup>f</sup> (n=1,188) | **78.6**<br>(221/281, 73.4–83.3) | **99.8**<br>(905/907, 99.2–100.0) | **99.1**<br>(221/223, 96.8–99.9) | **93.8**<br>(905/965, 92.1–95.2) |
| ≤ 7 days of symptom onset (n=929) | **81.9**<br>(199/243, 76.5–86.5) | **99.7**<br>(684/686, 99.0–100.0) | **99.0**<br>(199/201, 96.5–99.9) | **94.0**<br>(684/728, 92.0–95.6) |
| Asymptomatic Participants (n=877) | **68.8**<br>(33/48, 53.7–81.3) | **99.4**<br>(824/829, 98.6–99.8) | **86.8**<br>(33/38, 71.9–95.6) | **98.2**<br>(824/839, 97.1–99.0) |

<sup>a</sup>A three viral target RT-PCR assay was used. TaqPath SARS-CoV-2 Combo Kit.
https://www.fda.gov/media/136112/download
<sup>b</sup>Cycle threshold values are for the N gene (one of the three targets) for all specimens positive by RT-PCR.
<sup>c</sup>There were two indeterminate results from the initial BinaxNOW test and two indeterminate results from the repeated BinaxNOW test. All four were from separate enrollees, and the paired rapid test was negative except for one which had a threshold cycle of 30.7. Among discordant participants, 18/24 were symptomatic including all 14 participants with a negative to positive result.
<sup>d</sup>CI = Confidence Interval
<sup>e</sup>The repeated BinaxNOW test row defines positive as having either an initial or repeat BinaxNOW test that was positive at the same encounter.
<sup>f</sup>Defined as ≥ 1 current symptom from 15 symptom criteria for the COVID-19 case definition from the Council of State and Territorial Epidemiologists (CSTE). https://www.cste.org/news/520707/CSTE-Interim-Position-Statement-Update-to-COVID-19-Case-Definition.htm

Test positivity was 12.5% (265/2,110) for the initial BinaxNOW test, 12.7% (269/2,110) for repeat BinaxNOW test, and 15.8% (334/2,110) for RT-PCR. Of the 334 RT-PCR positive specimens, the N gene target Ct values for 258 specimens with positive initial BinaxNOW tests were significantly lower compared to 76 specimens with negative initial BinaxNOW results (20.4 vs 29.8, p<0.01, Supplementary Figure 1). Addition of a second BinaxNOW led to less than 1% increase in the combined percent positivity of BinaxNOW tests (279/2,110, 13.2%). The two sequential BinaxNOW tests were 98.9% concordant (2,086/2,110), with 255 concordant-positive and 1,831 concordant-negative pairs. There were 24 encounters with discordant BinaxNOW results (Table 1) of which 19 (79.2%) were RT-PCR positive.

The overall sensitivity of the initial BinaxNOW test compared to RT-PCR was 77.2% (258/334 95%CI 72.4–81.6), specificity was 99.6% (1,769/1,776, 95%CI 99.2– 99.8), PPV was 97.4% (258/265, 95%CI 94.6–98.9), and NPV was 95.8% (1,769/1,845, 95%CI 94.9–96.7) (Table 1). Among symptomatic individuals, the sensitivity of a single BinaxNOW test was 78.6% (221/281, 95%CI 73.4–83.3). In individuals within seven days of symptom onset, sensitivity was 81.9% (199/243, 95%CI 76.5–86.5). Among individuals reporting no current symptoms, the sensitivity was 68.8% (33/48, 95%CI 53.7–81.3). Repeating a second BinaxNOW test resulted in a sensitivity of 81.4% (272/334, 95%CI 76.8–85.5, Table 1). Asymptomatic antigen positive participants had a higher false positive proportion (5/38, 13.2%) compared to symptomatic antigen positive participants (2/223, 0.9%).

## Discussion

In this investigation of the BinaxNOW test, the overall sensitivity and sensitivity compared to RT-PCR among people with symptom onset within seven days were consistent with performance reported to the Federal Drug Administration (FDA) by the manufacturer [1]. Among asymptomatic individuals, sensitivity was lower consistent with other reports[2-4]. The reproducibility of the BinaxNOW test in this community setting by two separate groups of testers and individual readers was high. Only 1.1% of paired specimens tested within approximately 30 minutes of each other had differing BinaxNOW test results. When BinaxNOW tests were discordant, the RT-PCR was usually positive. Addition of a second BinaxNOW test at the same encounter did not significantly improve test sensitivity and offers low yield for capturing additional COVID-19 cases in this setting. Further work is needed to assess other antigen test combinations using a repeated test strategy and to identify the ideal frequency of repeat BinaxNOW testing.

This investigation supports foregoing confirmatory RT-PCR testing in symptomatic antigen test positive individuals per current guidelines, given <1% false positive BinaxNOW test results among symptomatic cases [9]. The FDA has warned healthcare workers of the potential for false positive antigen tests particularly in low-prevalence settings [10]. The BinaxNOW had overall high specificity regardless of symptom status (>99%) and high positive predictive value (97.4%) in a population with high prevalence (15.8%). Prevalence among asymptomatic individuals was lower (5.8%), but this population also had a relatively high positive predictive value (86.8%). In moderate-to-high pretest probability settings, foregoing a RT-PCR confirmatory test in asymptomatic antigen-positive individuals could be considered if resources are limited. The trade-off would be a relatively small but non-trivial increase in false positive results and the associated consequences (e.g., missed work/school, increased stress, unnecessary contact tracing).

False negative antigen tests are often consequential given the risk of furthering transmission due to perceived lack of infection. The significantly higher RT-PCR Ct values among BinaxNOW antigen-negative individuals suggest that nasal specimens from this group had less viral RNA (and possibly lower viral load) compared to antigen positive specimens. Others have shown reduced sensitivity of BinaxNOW antigen testing in asymptomatic individuals, and one explanation is that this group may be in the early infectious period or recovery period when viral load is lower [5, 11, 12].

There are several limitations to this investigation. The population was largely white, non-Hispanic, and older, and the findings may not be generalizable to other settings. Lack of swabbing supervision in other settings could alter test performance. Among asymptomatic individuals, a larger sample size is needed for more precise sensitivity estimates. RT-PCR was used as the gold standard for SARS-CoV-2 detection; however, since post-infectious individuals recovering from COVID-19 may have prolonged detectable viral shedding, the performance of BinaxNOW assays for detecting contagiousness may vary from these results. This investigation is specific to the BinaxNOW test, and the findings cannot be applied directly to other antigen tests.

This investigation provides BinaxNOW test performance in a real-world community setting. The BinaxNOW test exhibits minimal user error and high concordance (98.9%) when repeated at the same encounter offering low yield for capturing additional COVID-19 cases. Symptomatic BinaxNOW positive individuals in moderate-to-high pretest probability situations do not routinely need RT-PCR confirmatory tests because of high test specificity. In certain settings with high prevalence and limited resources, it may also be reasonable to forego RT-PCR confirmation in asymptomatic BinaxNOW positive individuals. Symptomatic negative individuals should continue isolation until RT-PCR confirmation. Quarantine should continue to be emphasized for asymptomatic BinaxNOW negative individuals after a close contact exposure. Given the costs and turn-around time for receiving RT-PCR results, antigen tests for SARS-CoV-2 are invaluable tools to break chains of transmission. Identification of the ideal frequency of antigen testing based on exposures, symptoms, RT-PCR turn-around times, and local incidence levels is needed.

## Data Availability

Data will be made available upon reasonable request.

## Acknowledgments

The authors thank the participants in Wisconsin for contributing to this investigation. This investigation was possible due to the vision of University of Wisconsin leadership including Tommy Thompson (President, University of Wisconsin System), Ray Cross (President Emeritus, University of Wisconsin System), Scott Neitzel (Vice President University Relations, University of Wisconsin System), Andrew J. Leavitt (Chancellor, University of Wisconsin Oshkosh), Stefan Fletcher (Director, Administrative Policies and Special Projects, University of Wisconsin System), Stacey Rolston (Interim Executive Director of UW-Shared Services, University of Wisconsin System), and Jo Carter (Project Portfolio Manager, University of Wisconsin System) as well as the U.S Department of Health and Human Services community-based testing program. The authors thank Ramika Archibald and Mary Wedig for assistance with reporting.

## Disclaimer

The findings and conclusions in this report are those of the author(s) and do not necessarily represent the official position of the Centers for Disease Control and Prevention (CDC). Use of trade names and commercial sources is for identification only and does not imply endorsement by the U.S. Department of Health and Human Services or CDC.

## References

1. US Department of Health and Human Services, Food and Drug Administration.BinaxNOW COVID-19 Ag Card. In Vitro Diagnostics Emergency Use Authorization. Silver Spring, MD. 2020. https://www.fda.gov/media/141567/download.

2. James AE, Gulley T, Kothari A, et al. Performance of the BinaxNOW COVID-19 Antigen Card test relative to the SARS-CoV-2 real-time reverse transcriptase polymerase chain reaction assay among symptomatic and asymptomatic healthcare employees. 2021: 1–15.

3. Okoye NC, Barker AP, Curtis K, et al. Performance Characteristics of BinaxNOW COVID-19 Antigen Card for Screening Asymptomatic Individuals in a University Setting. 2021.

4. Prince-Guerra JLJMM, report mw. Evaluation of Abbott BinaxNOW Rapid Antigen Test for SARS-CoV-2 Infection at Two Community-Based Testing Sites—Pima County, Arizona, November 3–17, 2020. 2021; 70.

5. Pilarowski G, Lebel P, Sunshine S, et al. Performance characteristics of a rapid SARS-CoV-2 antigen detection assay at a public plaza testing site in San Francisco. 2020.

6. UW–Madison offers surge testing to community amid rise in cases. November 11, 2020. https://news.wisc.edu/uw-madison-offers-surge-testing-to-community-amid-rise-in-cases/ Accessed February 1, 2021.

7. TaqPath™ COVID-19 Combo Kit. Food and Drug Administration. In vitro diagnostics EUAs. Silver Spring, MD: US Department of Health and Human Services, Food and Drug Administration; 2020.

8. Council of State and Territorial Epidemiologists (CSTE) Interim Position Statement: Update to COVID-19 Case Definition. August 7, 2020. https://www.cste.org/news/520707/CSTE-Interim-Position-Statement-Update-to-COVID-19-Case-Definition.htm Accessed February 1, 2021..

9. CDC. Coronavirus disease 2019 (COVID-19): interim guidance for antigen testing for SARS-CoV-2. Atlanta, GA: US Department of Health and Human Services, CDC; 2020. https://www.cdc.gov/coronavirus/2019-ncov/lab/resources/antigen-tests-guidelines.html

10. US Food and Drug Administration. Letter to Health Care Providers. Potential for False Positive Results with Antigen Tests, 11/3/2020. https://www.fda.gov/medical-devices/letters-health-care-providers/potential-false-positive-results-antigen-tests-rapid-detection-sars-cov-2-letter-clinical-laboratory Accessed February 1, 2021..

11. Perchetti GA, Huang M-L, Mills MG, Jerome KR, Greninger ALJJoCM. Analytical Sensitivity of the Abbott BinaxNOW COVID-19 Ag CARD. 2020.

12. Yu F, Yan L, Wang N, et al. Quantitative detection and viral load analysis of SARS-CoV-2 in infected patients. 2020; 71(15): 793–8.

