## Supplemental Information for "Performance of Repeat BinaxNOW SARS-CoV-2 Antigen Testing in a Community Setting, Wisconsin, November-December 2020"

**Supplementary Table 1:** Characteristics, symptoms, and initial Abbott’s BinaxNOW antigen test results for SARS-CoV-2 in persons providing nasal swabs (N = 2,110) – Oshkosh, Wisconsin, November 16 - December 15, 2020^a^

|  | **Overall**  **(n=2,110)** | **BinaxNOW Positive (n=265)** | **BinaxNOW Negative (n=1,845)** | **p-value** |
| --- | --- | --- | --- | --- |
| **Sex** |  |  |  | 0.45 |
| Female | 1,183 (56.1%) | 139 (52.5%) | 1,044 (56.6%) |  |
| Male | 905 (42.9%) | 123 (46.4%) | 782 (42.4%) |  |
| Unknown | 22 (1.0%) | 3 (1.1%) | 19 (1.0%) |  |
| **Age (years)** |  |  |  | 0.12 |
| 5 to 11 | 88 (4.2%) | 9 (3.4%) | 79 (4.3%) |  |
| 12 to 17 | 137 (6.5%) | 16 (6.0%) | 121 (6.6%) |  |
| 18 to 29 | 395 (18.7%) | 51 (19.2%) | 344 (18.6%) |  |
| 30 to 39 | 368 (17.4%) | 45 (17.0%) | 323 (17.5%) |  |
| 40 to 49 | 331 (15.7%) | 32 (12.1%) | 299 (16.2%) |  |
| 50 to 64 | 579 (27.4%) | 92 (34.7%) | 487 (26.4%) |  |
| 65 to 74 | 168 (8.0%) | 14 (5.3%) | 154 (8.3%) |  |
| 75 to 84 | 41 (1.9%) | 5 (1.9%) | 36 (2.0%) |  |
| ≥85 | 3 (0.1%) | 1 (0.4%) | 2 (0.1%) |  |
| **Ethnicity** |  |  |  | 0.37 |
| Non-Hispanic or Latino | 1,952 (92.5%) | 242 (91.3%) | 1,710 (92.7%) |  |
| Hispanic or Latino | 66 (3.1%) | 12 (4.5%) | 54 (2.9%) |  |
| Unknown | 92 (4.4%) | 11 (4.2%) | 81 (4.4%) |  |
| **Race** |  |  |  | 0.31 |
| White | 2,002 (94.9%) | 256 (96.6%) | 1,746 (94.6%) |  |
| Other^b^ | 62 (2.9%) | 5 (1.9%) | 57 (3.1%) |  |
| Multiple | 28 (1.3%) | 1 (0.4%) | 27 (1.5%) |  |
| Unknown | 18 (0.9%) | 3 (1.1%) | 15 (0.8%) |  |
| **Oshkosh City Resident**^c^ | 819 (38.8%) | 81 (30.6%) | 738 (40.0%) | 0.01 |
| **Healthcare Provider**^d^ | 190 (9.0%) | 22 (8.3%) | 168 (9.1%) | 0.91 |
| **University Affiliation**^e^ | 185 (8.8%) | 17 (6.4%) | 168 (9.1%) | 0.31 |
| **Close Contact in Past 14 Days** |  |  |  | <0.01 |
| Yes | 894 (42.4%) | 150 (56.6%) | 744 (40.3%) |  |
| No | 841 (39.9%) | 62 (23.4%) | 779 (42.2%) |  |
| Unknown | 375 (17.8%) | 53 (20.0%) | 322 (17.5%) |  |
| **Current Symptoms**^f^ |  |  |  | <0.01 |
| Yes | 1,188 (56.3%) | 223 (84.2%) | 965 (52.3%) |  |
| No | 877 (41.6%) | 38 (14.3%) | 839 (45.5%) |  |
| Unknown | 45 (2.1%) | 4 (1.5%) | 41 (2.2%) |  |
| **Current Symptoms** |  |  |  |  |
| Congestion/Rhinorrhea | 668 (31.7%) | 138 (52.1%) | 530 (28.7%) | <0.01 |
| Headache | 506 (24.0%) | 90 (34.0%) | 416 (22.5%) | <0.01 |
| Sore throat | 412 (19.5%) | 68 (25.7%) | 344 (18.6%) | <0.01 |
| Cough | 388 (18.4%) | 101 (38.1%) | 287 (15.6%) | <0.01 |
| Fatigue | 388 (18.4%) | 73 (27.5%) | 315 (17.1%) | <0.01 |
| Muscle Aches | 280 (13.3%) | 72 (27.2%) | 208 (11.3%) | <0.01 |
| Chills | 154 (7.3%) | 37 (14.0%) | 117 (6.3%) | <0.01 |
| Shortness of Breath | 127 (6.0%) | 19 (7.2%) | 108 (5.9%) | 0.48 |
| Loss of Smell | 106 (5.0%) | 61 (23.0%) | 45 (2.4%) | <0.01 |
| Diarrhea | 107 (5.0%) | 14 (5.3%) | 93 (5.0%) | 0.99 |
| Fever | 95 (4.5%) | 36 (13.6%) | 59 (3.2%) | <0.01 |
| Nausea/Vomiting | 95 (4.5%) | 13 (4.9%) | 82 (4.4%) | 0.86 |
| Loss of Taste | 92 (4.4%) | 48 (18.1%) | 44 (2.4%) | <0.01 |
| Abdominal Pain | 56 (2.7%) | 8 (3.0%) | 48 (2.6%) | 0.85 |
| Rigors | 1 (0.0%) | 0 | 1 (0.0%) | 1.0 |
| Other^g^ | 47 (2.2%) | 11 (4.2%) | 36 (2.0%) | 0.04 |

^a^This includes encounters where paired BinaxNOW tests and RT-PCR results were collected. Demographic data from 86 encounters were from repeat individuals.

^b^Includes Black, Asian, American Indian or Alaska Native, Native Hawaiian or Pacific Islander.

^c^Residency information was available for 2,109 encounters included in the denominator.

^d^Healthcare occupation information was available for 2,095 encounters included in the denominator.

^e^Includes faculty, students, and employees of a university and this data was available for 2,088 encounters included in the denominator.

^f^Defined as ≥ 1 current reported symptom from any of 15 symptom criteria for the COVID-19 case definition by the Council of State and Territorial Epidemiologists (CSTE). https://www.cste.org/news/520707/CSTE-Interim-Position-Statement-Update-to-COVID-19-Case-Definition.htm

^g^For encounters with none of the 15 listed CSTE symptoms reported, other symptoms included chest pain, throat swelling, ear pain, hoarse voice, and rash.

**Supplementary Figure 1:** Real-time reverse transcription–polymerase chain reaction (RT-PCR)^a^ SARS-CoV-2 threshold cycle values^b^ for 334 specimens collected in a community setting, stratified by the initial BinaxNOW antigen test status – Oshkosh, Wisconsin, November 16 - December 15, 2020
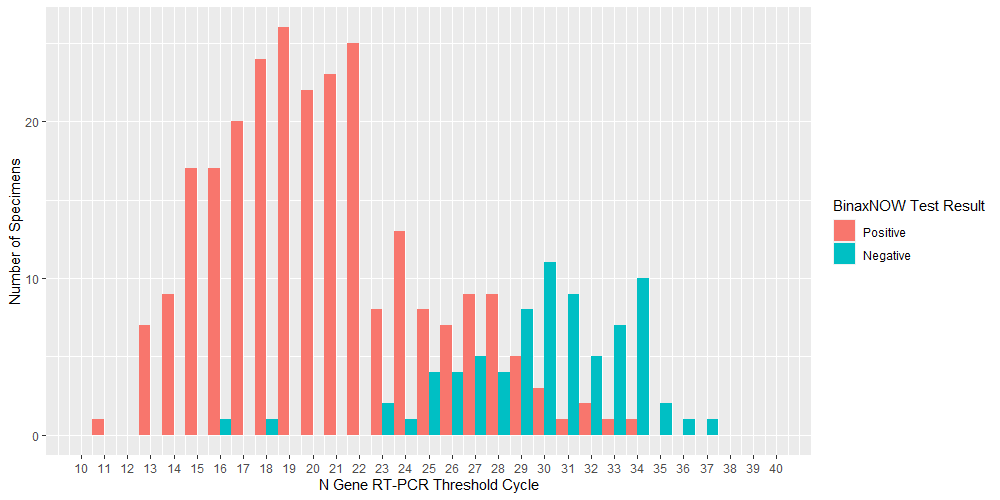

^a^A three viral target RT-PCR assay was used. TaqPath SARS-CoV-2 Combo Kit. https://www.fda.gov/media/136112/download

^b^For all specimens that met the criteria for RT-PCR positivity by this assay, we plotted the threshold cycle for the N gene (one of the three targets).

**Supplementary Figure 2:** Real-time reverse transcription–polymerase chain reaction (RT-PCR)^a^ SARS-CoV-2 threshold cycle values^b^ for 329 specimens collected in a community setting, stratified by symptom status^c^ – Oshkosh, Wisconsin, November 16 - December 15, 2020

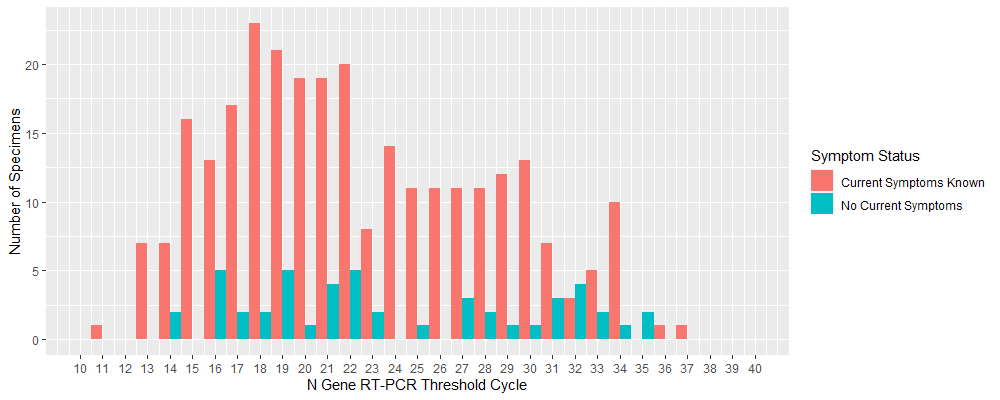

^a^A three viral target RT-PCR assay was used. TaqPath SARS-CoV-2 Combo Kit. https://www.fda.gov/media/136112/download

^b^For all specimens that met the criteria for RT-PCR positivity by this assay, we plotted the threshold cycle for the N gene (one of the three targets).

^c^Defined as ≥ 1 current reported symptom from any of 15 symptom criteria for the COVID-19 case definition by the Council of State and Territorial Epidemiologists. https://www.cste.org/news/520707/CSTE-Interim-Position-Statement-Update-to-COVID-19-Case-Definition.htm
